# Drinking motives and alcohol sensitivity mediate multi-dimensional genetic influences on alcohol use behaviors

**DOI:** 10.1101/2024.09.20.24314078

**Authors:** Jeanne E. Savage, Spit for Science Working Group, Danielle M. Dick, Danielle Posthuma

**Affiliations:** Department of Complex Trait Genetics, Centre for Neurogenomics and Cognitive Research, Amsterdam Neuroscience, Vrije Universiteit, Amsterdam, The Netherlands; Department of Psychiatry, Robert Wood Johnson Medical School, Rutgers Addiction Research Center, Rutgers University, Piscataway, NJ, USA; Department of Child and Adolescent Psychology and Psychiatry, section Complex Trait Genetics, Amsterdam Neuroscience, Vrije Universiteit Medical Center, Amsterdam, The Netherlands

**Keywords:** polygenic scores, level of response to alcohol, drinking motives, genetic heterogeneity, mediation

## Abstract

**Background:** Genetic influences account for a substantial proportion of individual differences in alcohol use behaviors (AUBs). However, multiple distinct sets of genes are linked to different AUBs, which may explain their dramatic variability in risk factors and manifestations. In this study, we explore whether intermediate neurobiological traits and alcohol-related cognitions mediate the relationship between polygenic scores (PGS) and multiple AUBs, with the aim to better understand processes captured by different genetic profiles.

**Methods:** Using results from prior genome-wide association studies, we derived PGS for 6 AUBs in participants from Spit for Science, a longitudinal study of college students in the U.S. (n=4,549). Self-report measures included personality traits, alcohol expectancies, drinking motivations, and alcohol sensitivity measures as well as drinking frequency, drinking quantity, alcohol use disorder (AUD) symptoms, and maximum drinks in 24 hours. Using linear regression and multiple mediation models, we investigated the direct and indirect effects of PGS on AUBs.

**Results:** In univariable regression results, PGSs indexing broad AUB dimensions such as drinks per week (DPW) and AUD predicted higher levels of sensation-seeking and multiple drinking motives, while *BeerPref* PGSs (indexing a variable pattern of alcohol problems associated with a preference for beer) predicted higher negative urgency and lower alcohol sensitivity. Mediational models indicated strong direct and indirect effects of DPW PGSs on multiple AUBs via social/enhancement drinking motives and alcohol sensitivity, indirect effects of AUD PGSs on AUD symptoms via coping motives, and indirect effects of *BeerPref* PGS on all AUBs via the joint effect of mediators including alcohol sensitivity.

**Conclusions:** These findings provide initial evidence that the genetic influences on different AUBs are associated with and partially mediated by intermediate neurobiological and cognitive factors, which may be more amenable to intervention. Greater focus on drinking motives and alcohol sensitivity is warranted in genetic research, as well as attention to the heterogeneous pathways linking genes to alcohol use outcomes.

## Introduction

Alcohol use behaviors (AUBs) manifest in a variety of ways, with substantial individual and population differences in the frequency, quantity, timing, and types of alcohol that people consume (if they choose to drink at all) (Litten et al., 2015, Rehm et al., 2003).

AUBs also vary substantially across environments and throughout the lifespan. For example, individuals in Mediterranean countries typically consume moderate amounts of alcohol (primarily wine) and drink frequently but almost exclusively with meals, while individuals in northern and eastern European countries drink to intoxication more often and tend to drink beer and spirits (Sieri et al., 2002, Simpura and Karlsson, 2001). Young adults typically drink on fewer days but engage in higher levels of heavy episodic and risky drinking behaviors, while older adults consume smaller quantities of alcohol more frequently (Leggat et al., 2022, Britton et al., 2015, Holton et al., 2019). Clinically significant alcohol use disorders (AUD) are marked by a dynamic progression towards heavier, uncontrolled drinking, although the tempo and intensity of this trajectory also differs between individuals.

The factors influencing AUBs are as varied as the behaviors themselves. At the most basic level, genes have a robust impact on AUBs, with heritability estimates of 50-60% for drinking frequency, quantity, maximum drinks in 24 hours, alcohol problems, and lifetime AUD diagnoses (Agrawal et al., 2012, Dick et al., 2011, Verhulst et al., 2015). The effect sizes of individual genes or single nucleotide polymorphisms (SNPs) in the DNA are generally extremely small. Intermediate neurobiological traits such as personality, brain structure, and reward sensitivity correlate modestly, but consistently, with AUBs (Gunn et al., 2013, Whelan et al., 2014). More proximally, alcohol-related cognitions, such as expectancies about the positive and negative effects of alcohol and motivations for drinking, as well as alcohol-specific metabolic processes (alcohol sensitivity) have robust and direct associations with AUBs (Bresin and Mekawi, 2021, Schuckit et al., 1997, Agrawal et al., 2008, Schuckit, 2009). There is some evidence that the genetic influences on AUBs are partially mediated by these intermediate and proximal factors (Littlefield et al., 2011, Prescott et al., 2004, Li et al., 2017, Kendler et al., 2021). However, such mediational studies are few, and most apply biometrical analyses to infer genetic influences without providing insight into molecular genetic functions. They also focus on parsing the genetic influences on only a few alcohol measures, usually lumping these into broad dimensions such as overall consumption levels or lifetime AUD diagnoses.

Emerging evidence from genome-wide association studies (GWASs), however, has demonstrated that the relationships amongst AUBs are complex. While “alcohol consumption” and “alcohol problems” form consistent, coherent dimensions, they are genetically distinct from each other (Deak and Johnson, 2021, Sanchez-Roige et al., 2019). There are also important unique genetic influences on drinking frequency versus quantity (Mallard et al., 2022), and on patterns of drinking and beverage preference (Savage et al., 2023). By the time these genetic effects are diluted through a chain of biological processes and moderated by external factors on their pathway to influencing AUBs, it’s plausible that etiological distinctions between dimensions may be even greater.

The heterogeneity of AUBs and their underlying causes poses a challenge for efforts to develop personally tailored prevention and intervention efforts (Litten et al., 2015).

Personalized strategies can be more effective than universal programs for reducing risky or harmful AUBs (Cronce and Larimer, 2011, Savage et al., 2015), which is especially critical as prevention programs often have small effects on reducing alcohol use (Strøm et al., 2014) and relapse is a prominent feature of AUD treatment (Sliedrecht et al., 2019). However, proper targeting of interventions requires an understanding of the etiology of a behavior for each specific individual. Incorporating genetic predictions, in the form of polygenic risk scores (PGSs), is an ambition for many such personalized medicine applications (Lewis and Vassos, 2020) as DNA has desirable qualities for an intervention target. One’s genetic code is not subject to reverse causation, so the direction of effect is clear; DNA is fixed before birth, so it is possible to identify at-risk individuals prior to the onset of problems; and one’s genome provides the ultimate level of individual-specific prediction (save for monozygotic twins). Yet genetic instruments are not commonly used in personalized interventions for AUBs, in no small part due to the fact that our current knowledge of the links between DNA variation and AUBs provides very poor individual-level accuracy (<5%) in disease/phenotype prediction (Saunders et al., 2022, Zhou et al., 2023). However, this low accuracy may itself be a result of heterogeneity in the phenotype definition. As both gene discovery and PGS validation efforts are based almost exclusively on broad consumption and AUD phenotypes, genetic effects specific to different AUBs will be washed out, even as larger discovery sample sizes allow for the identification of more and more associated genes with smaller and smaller effects (Deak and Johnson, 2021).

One solution to this challenge is to focus gene identification efforts on more diverse and refined measures of AUBs (Wong and Schumann, 2008, Mallard et al., 2022, Savage et al., 2023), which provide better insight into their genetic architecture (although it requires much more effort and a higher participant burden to collect such measures at scale). But beyond this, PGSs can be applied not only for direct prediction of heterogeneous consumption/AUD measures, but also to investigate an array of candidate intermediate processes to better understand the varied mechanisms connecting genes to AUBs (Li et al., 2017, Salvatore et al., 2014). For the most part, the many genes that have been linked to AUBs show only a statistical association, and their etiological processes are, as yet, unknown (Deak and Johnson, 2021). However, predicting that someone is at higher or lower genetic risk is not as helpful as knowing *why* they might be at risk – the actual process that could be intervened upon. In combination with more refined gene discovery efforts, PGSs have the potential to identify more proximal processes, such as alcohol expectancies and drinking motives, through which genes exert their effects. Such constructs are much more amenable to manipulation than the DNA itself and have been demonstrated to be effective targets for intervention (Hingson, 2010). By linking genetic factors to precise intermediate processes, a greater level of specificity could also be achieved for developing individually tailored interventions. Knowing which prevention or treatment is best suited to an individual could improve outcomes and reduce the amount of time spent in unsuccessful treatment attempts, along with negative health and societal consequences during those periods.

In this study, we investigate the intermediate mechanisms underlying genetic influences on multiple dimensions of AUBs. Applying a multiple mediation model, we examine the parallel role of drinking motives, alcohol expectancies, alcohol sensitivity, and personality traits linking individual differences in the genome to individual differences in AUBs. These findings have the potential to provide better insight into the statistical associations identified in GWAS as well as guiding future applications of interventions based on individual-level genetic risk.

## Materials and Methods

### Participants

Data was derived from “Spit for Science” (S4S), a prospective longitudinal study of over 12,000 college students at a large, urban, public university (Dick et al., 2014). Across multiple incoming student cohorts, all first-time freshmen aged 18+ were eligible to complete a self-report survey and provide a saliva sample for DNA collection early in the fall of their first year (Y1F). Follow-up surveys were sent out each subsequent spring to participants still enrolled at the university (Y1S-Y4S). Participants were 62.7% (cis gender) female and 48.6% self-reported their race and ethnicity as White. All participants provided informed consent and the S4S study was approved by the university Institutional Review Board.

After applying inclusion criteria (below), the current study used data from *N* = 4,549 S4S participants from the first 5 cohorts for whom genotyping was complete. Enrollment rates were high, with 64% of the eligible incoming students completing an initial survey and 42%-73% returning to complete follow-up surveys across the subsequent data collection waves. Nearly all participants, 97%, provided a DNA sample. Included participants were restricted to those whose genomes were most similar to European ancestry reference panels and whose DNA samples passed genetic quality control thresholds (Webb et al., 2017, Dick et al., 2014, Peterson et al., 2017). Ancestry filtering was applied due to the need to match ancestral background to the European discovery GWAS sample, as there is poor genetic prediction possible across ancestry groups due to inherited differences in patterns of linkage disequilibrium (LD) in the genome.

### Measures

Eligible participants were emailed a link to complete a confidential online self-report survey which assessed a wide range of traits and behaviors, with a focus on alcohol use. Data was collected and managed by the secure, web-based REDCap system of electronic data capture tools (Harris et al., 2009). Measures were largely derived from psychometrically validated scales administered in an abbreviated version or staggered across waves/cohorts to reduce participant burden. Psychometric properties and further details of most scales in this sample have been published previously (Dick et al., 2014, Savage and Dick, 2023a, Salvatore et al., 2016, Savage et al., 2022). Descriptive statistics for each of the measures are shown in **Table 1**. Stable traits such as personality were assessed in the initial surveys (Y1F and Y1S), while alcohol-related measures were collected at every wave. For cohort 5, only data through Y2S was used due to the onset of the Covid-19 pandemic in Y3S and subsequent changes in both the campus environment and study protocols.

**Table 1.**
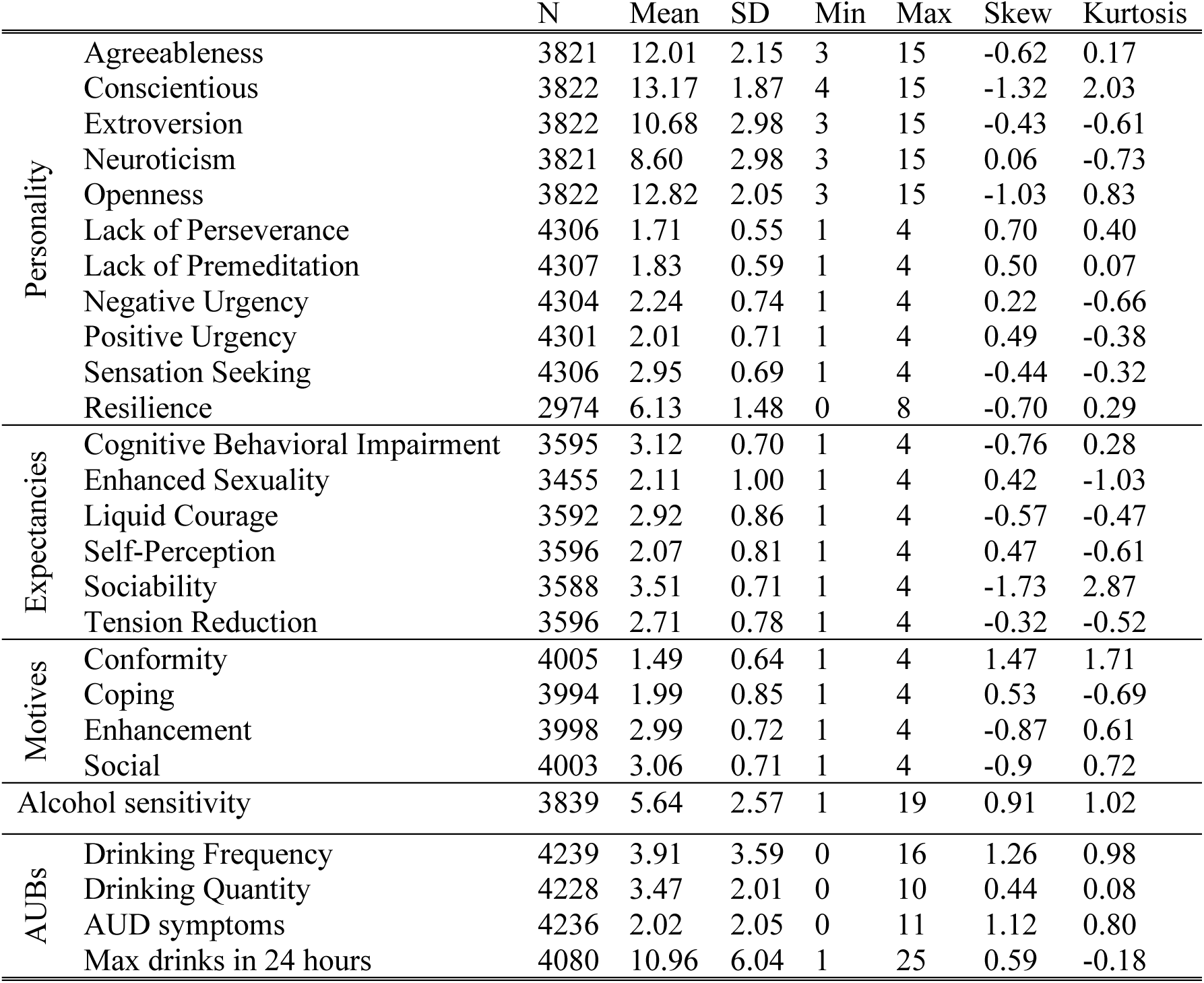
Descriptive statistics of alcohol use behavior (AUB) measures and mediators.

#### Personality Traits

The surveys assessed personality traits using a subset of items for each subscale of the Big Five Inventory (BFI; John and Srivastava, 1999), each impulsivity-related subscale (sensation-seeking, negative urgency, positive urgency, lack of premeditation, lack of perseverance) of the UPPS-P (Lynam et al., 2006), and the Connor-Davidson resilience scale (CD-RISC; Connor and Davidson, 2003). Prorated sum scores were available for the BFI subscales in cohorts 1-4. Mean scores were available for the UPPS-P subscales for all cohorts and for the CD-RISC scale in cohorts 1-3.

#### Alcohol Expectancies

In the freshman and sophomore year waves (Y1F-Y2S), participants were asked about what effects they expected to experience from drinking alcohol (whether or not they had yet initiated use). These included 6 subscales of Cognitive-Behavioral Impairment, Enhanced Sexuality, Liquid Courage, Sociability, Tension Reduction, and Self-Perception from the Alcohol Expectancies Questionnaire (AEQ; Fromme et al., 1993). Subscale scores were averaged across waves to create a single mean score.

#### Drinking Motives

In each survey, participants who had initiated drinking completed an abbreviated version of the Drinking Motives Questionnaire – Revised (Cooper, 1994).

Mean scores were derived for four subscales: Coping, Enhancement, Conformity, and Social motives. Subscale scores were averaged across waves.

#### Alcohol Sensitivity

In each survey, participants who reported having used alcohol 5 or more times in their lives (85.8%) responded to the Self-Rating of the Effects of Alcohol (SRE) scale (Schuckit et al., 1997). This scale consists of 4 questions that ask students to think back to the first 5 times they consumed alcohol and report how many standard drinks it took for them to feel tipsy/have a buzz, feel dizzy/slur their speech, stumble/find it hard to walk, and fall asleep without intending to. The average number of drinks needed to feel these intoxicating effects was used to define the SRE, with lower values reflecting a higher alcohol sensitivity. SRE scores from the first available self-report were used for analysis to ensure that recall was as close in time as possible to the initiation of alcohol use.

#### Alcohol Use Behaviors

At each wave, participants were asked if they had initiated alcohol use, and, if so, about a variety of AUBs. These included questions about typical drinking frequency (number of drinking days per month; Freq) and typical drinking quantity (number of drinks per drinking day; Quant) from the AUDIT questionnaire (Bohn et al., 1995), *DSM-5* AUD symptoms (AUDsx) from the Semi-Structured Assessment for the Genetics of Alcoholism (SSAGA; Bucholz et al., 1994), and maximum drinks consumed in a 24-hour period in the past year (Max24), also from the SSAGA. Frequency and quantity measures were recoded to a pseudo-continuous number of days per month or drinks per day using the mean of each range category (Salvatore et al., 2016). Mean values of Freq, Quant, and AUDsx across waves, and maximum values of Max24, were used for analysis.

### Polygenic scores (PGSs)

PGSs were derived from summary statistics of a prior genomic structural equation modelling GWAS in the UK Biobank sample (Savage et al., 2023), which identified four latent genetic factors underlying a set of 18 normative and problematic AUBs. These factors represented (1) chronic and severe alcohol *Problems*, (2) *BeerPref*, a decreasing pattern of alcohol use/problems later in life marked by a preference for drinking beer and drinking without accompanying meals, (3) overall quantity and frequency of *Consumption* of varied alcoholic beverage types, and (4) *AtypicalPref*, a preference for drinking less common types of beverages such as fortified wine and spirits. GWAS of these factors was conducted in up to 386,961 individuals from the UK Biobank sample, representing the largest genetic investigation to date of several specific AUB dimensions. However, much larger GWASs have been carried out on broad AUB measures such as typical drinking quantity and AUD diagnoses. As a means of comparing whether specific AUB measures add information over and above these more powerful (but likely more heterogeneous) measures, we additionally examined PGSs for the largest available GWAS of drinks per week (DPW; Saunders et al., 2022) and AUD (Zhou et al., 2020).

For each of these 6 measures, we created PGSs based on previously published GWAS summary statistics. For DPW and AUD, we used results excluding UK Biobank to avoid overlap with the four latent AUB factor measures. GWAS summary statistics were weighted using PRS-CS “auto” version and European LD reference panels from UK Biobank (four latent factors) or 1000 Genomes (DPW; AUD), as provided with the software (Ge et al., 2019). PGS were calculated in PLINK2 (Chang et al., 2015) using the --score method. SNPs in the S4S dataset were first filtered on imputation INFO score > .8, minor allele frequency >.05, Hardy-Weinberg equilibrium *p* values > 5x10^-6^, and missingness < .025. Full details about genotyping and quality control procedures for this sample have been described elsewhere (Webb et al., 2017, Dick et al., 2014). Each PGS was entered in a regression model with 10 ancestry principal components, genetic sex, and age (mean across available data waves) as predictors. The PGS residuals, after removing the effects of these covariates, were used for further analysis.

### Data Analysis

All measures were standardized prior to analysis. First, we examined the overlap of genetic information between the various PGSs by calculating pairwise Pearson correlations between each PGS. Second, we conducted a series of univariable linear regression analyses to identify which AUBs and potential mediators demonstrated evidence of association with each PGS. These analyses were carried out with the lm() function in R version 4.2.2 (R Core Team, 2017). Nominally significant associations (*p* < .05) between each PGS and measures of personality, alcohol expectancies, drinking motives, or alcohol sensitivity were selected for the full model. Finally, these selected associations were included as mediational paths linking PGSs from the four latent AUB factors to each measured AUB in a multiple mediation model (**Figure 1**), while accounting for the correlations between these variables. A similar model was estimated using the PGSs of DPW and AUD as a comparison. Mediational analyses were carried out with structural equation modelling in Mplus version 8.4 (Muthén and Muthén, 2011), using robust maximum likelihood estimation to account for missing and non-normally distributed data. The models provided estimates of the effect of each PGS on each AUB outcome, both from direct paths and indirect effects via the mediators. Bonferroni correction for 6 PGSs and 4 AUB outcome measures was applied (.05/(6*8) = .002) to evaluate the significance of these direct and indirect effects.

**Figure 1.**
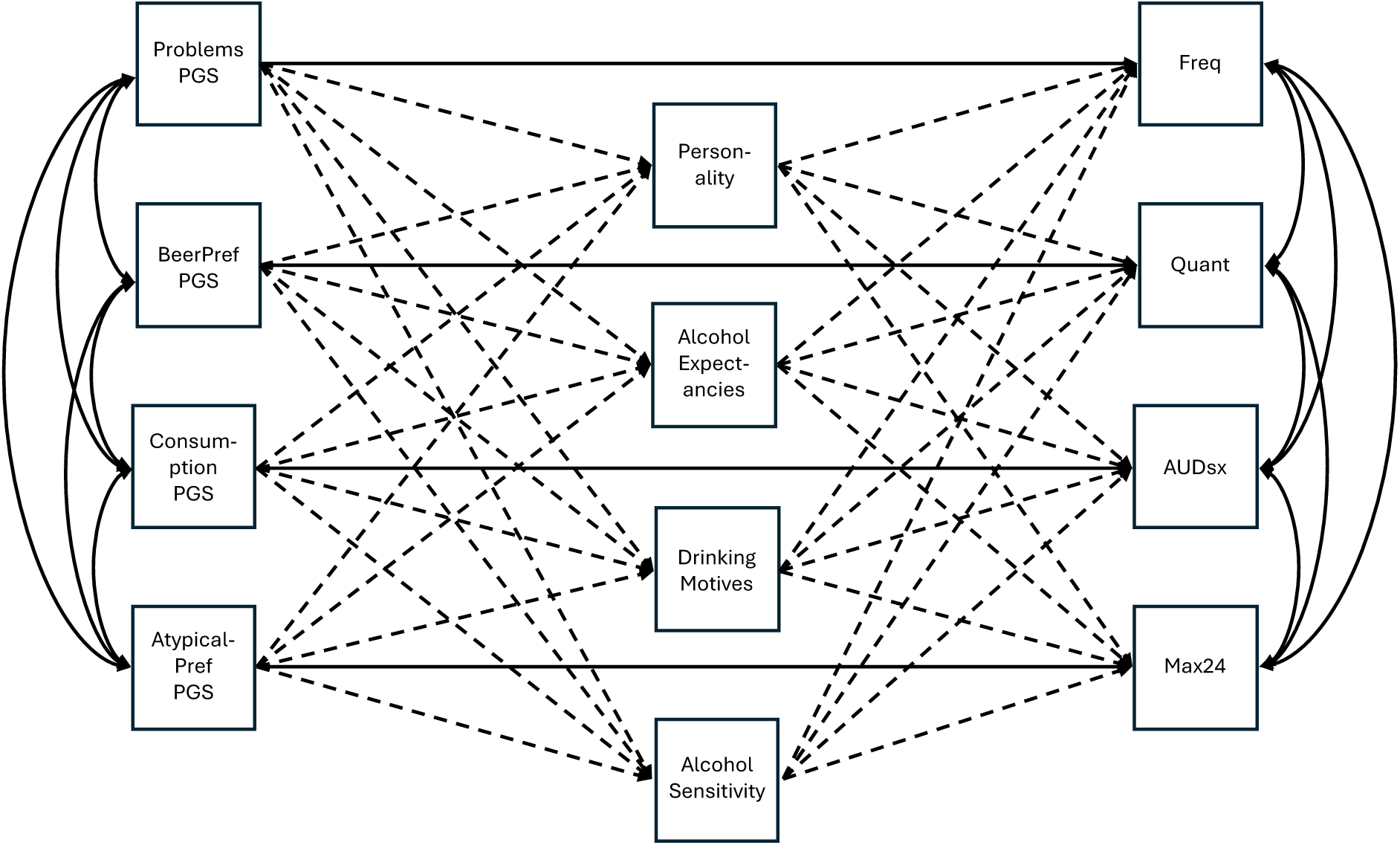
Example diagram of the multiple mediation model linking polygenic scores to alcohol use behaviors. Solid lines represent direct effects and dashed lines indirect effects. Single-headed arrows represent directional association pathways and double-headed arrows represent correlations.

## Results

Correlations between the 6 PGSs used ranged from -.096 (BeerPref - Consumption) to .393 (Problems - Consumption), reflecting the heterogeneity of genetic influences captured by these different AUB dimensions (**Table 2**). Univariable regression results demonstrated that most AUBs and potential mediators had a nominal association with at least one PGS, although the patterns were varied (**Table 3**). *Problems* PGSs predicted higher levels of negative and positive urgency, Freq, and Max24. *BeerPref* predicted higher extroversion, lower expectations of cognitive-behavioral impairment from alcohol, lower alcohol sensitivity, lower conformity motives, and higher Quant, AUDsx, and Max24. *Consumption* predicted lower conscientiousness and extroversion and higher lack of perseverance, enhancement and social motives, and Freq. *AtypicalPref* was associated only with lower conformity motives and a lower alcohol sensitivity. DPW and AUD were both highly significantly associated with higher levels of all AUBs and drinking motives (only AUD with conformity motives), as well as multiple impulsivity traits and alcohol expectancies. A few dimension-specific associations were also observed, such as between DPW and higher lack of perseverance, and between AUD and higher negative urgency. Perhaps surprisingly, expectations of tension reduction effects from alcohol were not linked to any AUB PGS.

**Table 2.** Genetic correlations between polygenic scores for alcohol use behaviors.

|  | 1 | 2 | 3 | 4 | 5 | 6 |
| --- | --- | --- | --- | --- | --- | --- |
| 1. Problems | 1.000 |  |  |  |  |  |
| 2. BeerPref | 0.381 | 1.000 |  |  |  |  |
| 3. Consumption | 0.393 | -0.096 | 1.000 |  |  |  |
| 4. AtypicalPref | 0.254 | 0.259 | 0.132 | 1.000 |  |  |
| 5. DPW | 0.238 | 0.036 | 0.338 | 0.077 | 1.000 |  |
| 6. AUD | 0.329 | 0.215 | 0.234 | 0.096 | 0.276 | 1.000 |

*Note: DPW=drinks per week; AUD=alcohol use disorder.*

**Table 3.**
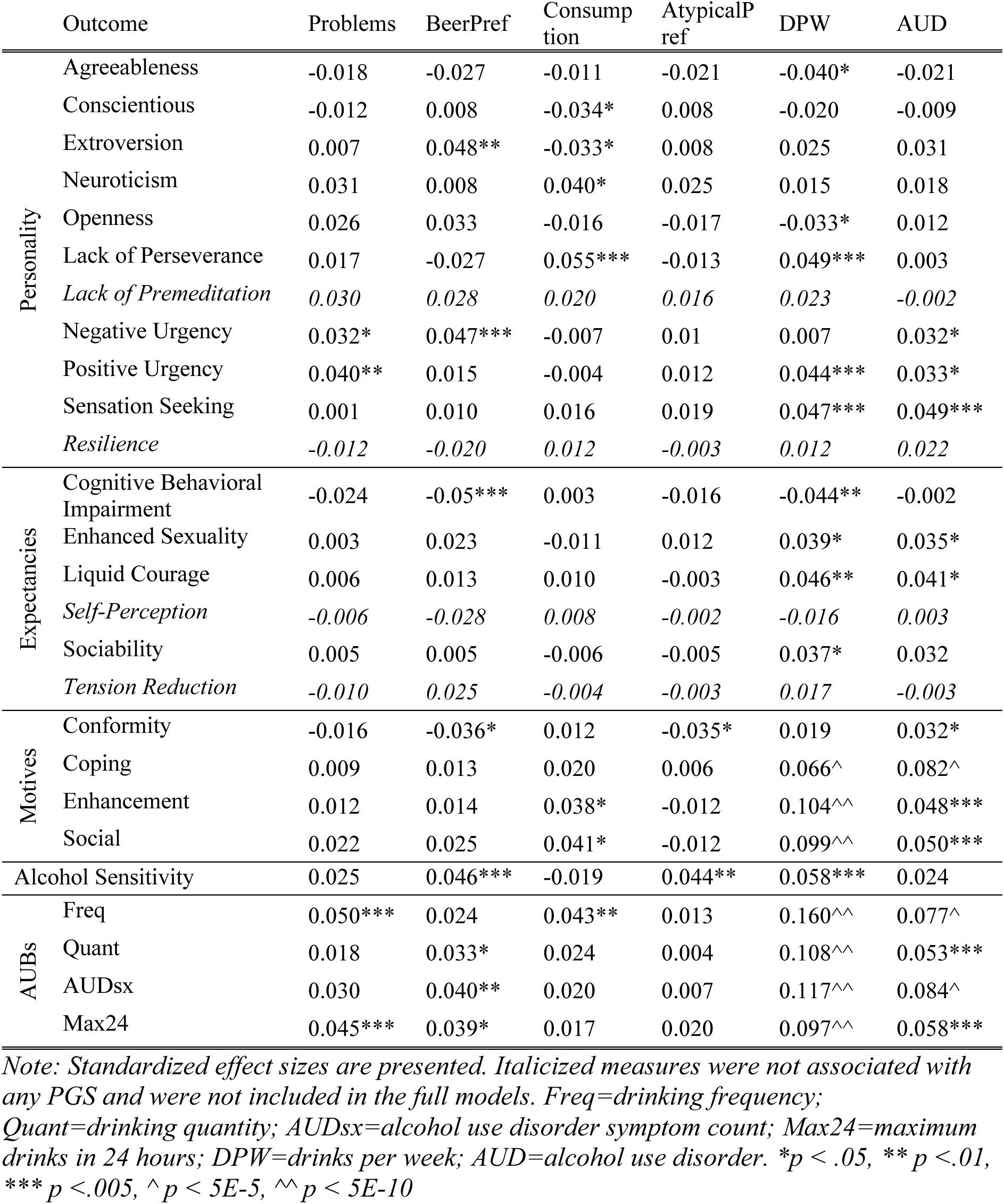
Univariable regression associations between polygenic scores (PGS) and alcohol use behavior (AUB) measures and mediators.

Nominal associations from the univariable regressions were carried forward to multiple mediation models. In the first analysis, focused on the latent AUB factor PGSs, the full model explained between 12.4% and 36.5% of the variance in each of the measured AUBs (**Table 4**). However, this was mostly driven by the mediators themselves, as the direct and indirect effects of the PGSs were small (maximum standardized ß = .043). Only the *BeerPref* PGS had significant associations in this model after multiple testing correction, showing significant indirect effects (total standardized ß = .008-.039) on higher levels of each of the measured AUBs. The largest share of these came from intermediate effects on a lower alcohol sensitivity (standardized ß =.005-.021), although the specific indirect pathways were not statistically distinguishable from each other.

**Table 4.**
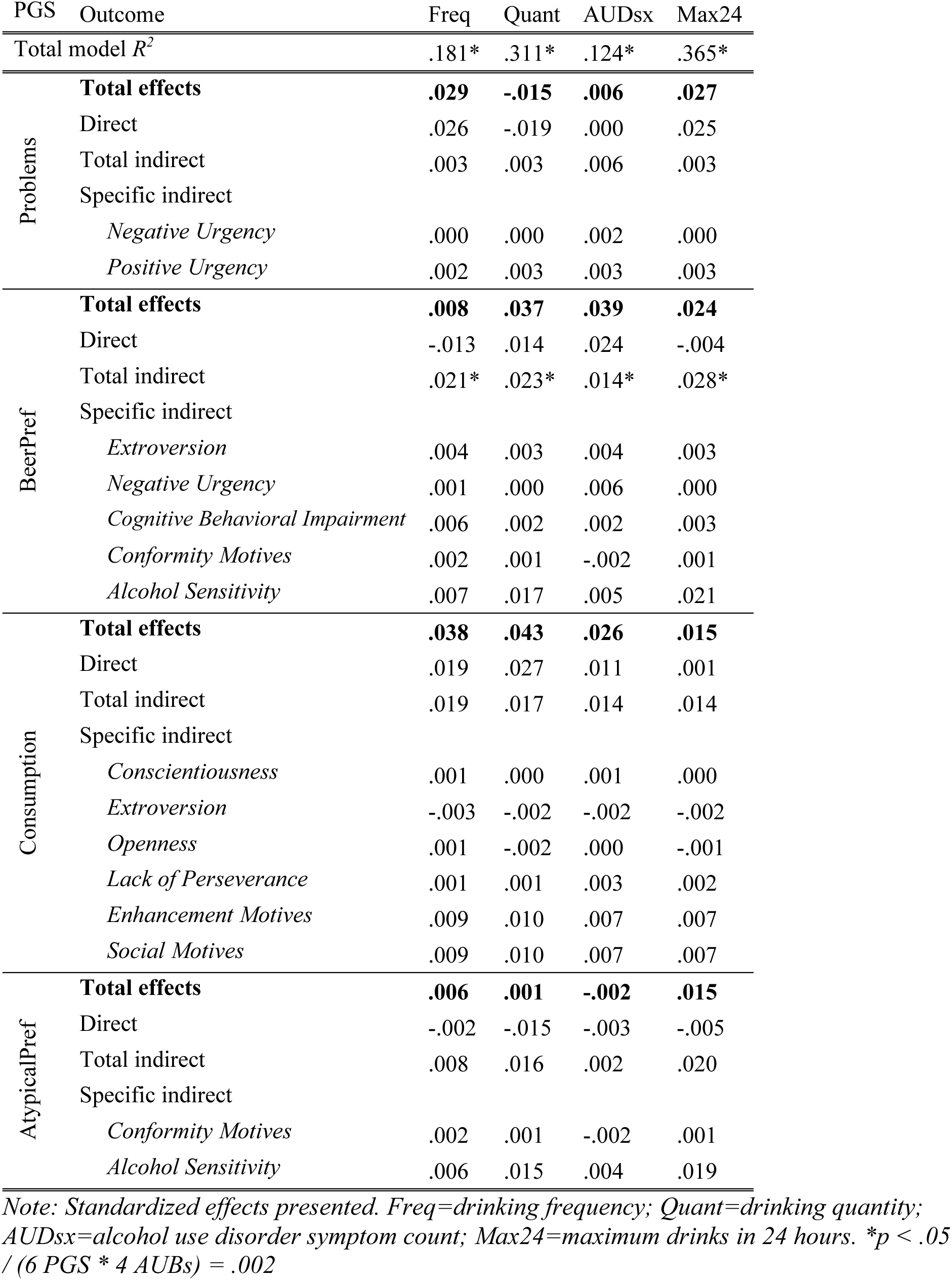
Direct and indirect effects of polygenic scores for latent factors of alcohol use behavior (AUB) measures predicting AUBs in multiple mediation models.

Results from the second model, focused on the higher-powered PGSs of DPW and AUD, are shown in **Table 5**. In this model, 13.9% to 35.6% of the variance in each of the measured AUBs was explained, in this case largely by the genetic scores. DPW PGS had significant direct effects (standardized ß = .060-.094) on Freq and AUDsx, and indirect effects on all AUBs (standardized ß = .043-.081), which were specifically mediated through higher levels of positive reinforcement drinking motives (enhancement and social) and a lower level of alcohol sensitivity. AUD PGSs, on the other hand, only had significant total indirect effects on AUDsx that were largely mediated through coping motives, as well as significant specific indirect effects on Freq and Max24 through higher coping motives.

**Table 5.** Direct and indirect effects of polygenic scores for DPW and AUD predicting alcohol use behavior (AUB) measures in multiple mediation models.

| PGS | Outcome | Freq | Quant | AUDsx | Max24 |
| --- | --- | --- | --- | --- | --- |
| Total $R^2$ | | .181 | .297 | .139 | .356 |
| DPW | **Total effects** | **.155\*** | **.112\*** | **.103\*** | **.089\*** |
|  | Direct | .094* | .031 | .060* | .012 |
|  | Total indirect | .062* | .081* | .043* | .078* |
|  | Specific indirect |  |  |  |  |
|  | *Agreeability* | .001 | .001 | .000 | .002 |
|  | *Neuroticism* | .001 | .002 | .000 | .002 |
|  | *Lack of Perseverance* | .002 | .001 | .003 | .002 |
|  | *Positive Urgency* | .001 | .002 | .002 | .001 |
|  | *Sensation Seeking* | .003 | .003 | .001 | .004 |
|  | *Cognitive Behavioral Impairment* | .005 | .002 | .003 | .003 |
|  | *Liquid Courage* | .002 | .002 | .005 | .002 |
|  | *Sexuality* | .000 | .000 | .002 | .000 |
|  | *Sociability* | .000 | .002 | .001 | .001 |
|  | *Conformity Motives* | -.001 | .000 | .000 | .000 |
|  | *Enhancement Motives* | .018* | .021* | .009* | .014* |
|  | *Social Motives* | .019* | .023* | .011* | .016* |
|  | *Alcohol Sensitivity* | .010* | .024* | .007* | .032* |
| AUD | **Total effects** | **.039** | **.024** | **.061\*** | **.027** |
|  | Direct | .018 | .008 | .027 | .008 |
|  | Total indirect | .022 | .016 | .034* | .018 |
|  | Specific indirect |  |  |  |  |
|  | *Negative Urgency* | .001 | .001 | .003 | .000 |
|  | *Positive Urgency* | .001 | .001 | .002 | .001 |
|  | *Sensation Seeking* | .004 | .003 | .002 | .004 |
|  | *Liquid Courage* | .001 | .002 | .004 | .002 |
|  | *Sexuality* | .000 | .000 | .001 | .000 |
|  | *Conformity Motives* | -.002 | -.001 | .001 | -.001 |
|  | *Coping Motives* | .008* | .000 | .016* | .005* |
|  | *Enhancement Motives* | .005 | .006 | .002 | .004 |
|  | *Social Motives* | .004 | .005 | .002 | .003 |

Note: Standardized effects presented. Freq=drinking frequency; Quant=drinking quantity; AUDsx=alcohol use disorder symptom count; Max24=maximum drinks in 24 hours; DPW=drinks per week; AUD=alcohol use disorder. \* $p < .05 / (6 \text{ PGS} * 4 \text{ AUBs}) = .002$

## Discussion

Combining large-scale gene discovery efforts with mediational modelling in a deeply phenotyped sample, this study has demonstrated that aggregated genetic variants linked to AUBs are also associated with a variety of plausible intermediate mechanisms that influence individual differences in AUBs. Evidence supported a role of drinking motives mediating pathways between genes and broad dimensions of AUBs, including coping motives accounting for a large portion of the identified genetic risk for AUD symptoms and social and enhancement motives significantly linking DPW-related genes to multiple AUBs. The effect of DPW-associated genes was also strongly mediated by a low level of sensitivity to alcohol. Furthermore, some evidence supported the importance of specific genetic dimensions of AUBs. Genes previously associated with preference for beer and a pattern of transient consumption/problems (*BeerPref*) were also linked indirectly to AUBs via intermediate pathways such as low alcohol sensitivity. By simultaneously examining the role of multiple sets of genes and intermediate traits, we were able to account for a relatively high proportion of individual variation in AUBs as well as gain insight into the mechanisms by which multiple dimensions of genetic susceptibility manifest.

These results continue to support the notion that AUBs, and their underlying causes, are heterogeneous (Litten et al., 2015, Mallard et al., 2022, Wong and Schumann, 2008).

First, genetic scores indexing multiple AUB dimensions were only modestly correlated with each other. Second, these PGSs demonstrated unique patterns of associations with intermediate measures in the univariable regression models. While these in part likely reflect power differences from the GWAS discovery samples, there were also some qualitative differences in the patterns observed. For example, while *Consumption* and DPW are conceptually similar measures of consumption behavior, they exhibited opposite patterns of association with mediators such as extroversion, positive urgency, expectancies of cognitive behavioral impairment, and alcohol sensitivity. For these measures, the pattern of associations for DPW PGS seemed more similar to those of the *BeerPref* PGS, suggesting that the pathways captured by the broader DPW measure may contain a mix of genetic influences on dynamic (*BeerPref*) versus persistent (*Consumption*) drinking behaviors. It is also notable that none of the 4 latent factor AUB PGSs had even marginal associations with sensation seeking, a robust risk factor for AUBs (Adams et al., 2012, Li et al., 2017, Stautz and Cooper, 2013) which was strongly associated with both DPW and AUD PGSs in the univariable analyses in this sample. It is plausible that the genes indexed by these AUB domains are more specific to alcohol-related processes, while the broader DPW and AUD domains capture more influences of a general externalizing/reward seeking predisposition that is robustly, but non-specifically, linked to AUBs – as well as other substance use behaviors and impulsivity traits (Kendler et al., 2011, Poore et al., 2023).

Heterogeneity was also observed in the patterns of associations between PGSs and AUBs in the mediational models. DPW PGSs were associated with all AUBs but most strongly with drinking frequency (especially in the direct genetic effects), despite drinks per week in fact being a measure of quantity rather than frequency. Previous research has shown that genetic influences on total consumption measures like DPW can be biased by external factors such as socioeconomic status, which have specific effects on drinking frequency rather than quantity (Mallard et al., 2022). AUD PGSs, on the other hand, were more specifically associated with AUDsx. *BeerPref* had indirect effects on all AUBs, with the strongest total indirect effects on drinking quantity and AUDsx. The genetic influences on this factor might be relevant to age or developmental differences in the processes underlying AUBs since this factor represents patterns of longitudinal changes in consumption as well as alcohol problems earlier rather than later in life (i.e. individuals who may age out of drinking/drinking problems). Our results indicate that these influences are especially relevant to AUBs in this young adult cohort. Such dynamics are not well characterized in typical GWASs of static measures such as current DPW in (older) adults or a dichotomous lifetime AUD diagnosis. This factor is especially interesting because its dynamic nature may represent biological mechanisms that facilitate recovery or are more sensitive to intervention.

Of the intermediate mechanisms investigated, the strongest evidence for mediation of genetic effects was observed for alcohol sensitivity and drinking motives. This is consistent with a large body of evidence demonstrating that these factors have robust, proximal effects on AUBs (Schuckit, 2009, Kuntsche et al., 2005, Bresin and Mekawi, 2021). However, there have been relatively few investigations of the etiology of these constructs using measured genes (Savage et al., 2022, Lai et al., 2020) rather than twin studies (which are not informative about the specific genes involved), and even fewer establishing whether their genetic influences overlap those of AUBs. Here, we demonstrate that many of the genetic variants linked to individual differences in AUBs appear to exert their influence via indirect pathways including alcohol sensitivity and motivations for drinking. Further, consistent with epidemiological literature (Bresin and Mekawi, 2021, Savage and Dick, 2023b), positive reinforcement social and enhancement motives mediate genetic risk for (heavy) consumption while negative reinforcement coping motives mediate AUD risk genes, indicative of multiple distinct etiological processes. It is also interesting that the genetic influences on *BeerPref* were strongly associated with and partially mediated by alcohol sensitivity. Unlike most AUBs, *BeerPref* is not associated with genetic variation in the regions of the genome containing alcohol metabolism genes in the *ADH* and *ALDH* gene families (Savage et al., 2023). This suggests an important biological effect on the subjective feelings of intoxication that may be independent of direct alcohol metabolic processes, yet relevant to downstream AUBs. More biobehavioral research on the genetic etiology of these intermediate constructs is needed enhance future therapeutic applications, for example, to reduce the rewarding effects of alcohol or dampen motivational drives for consumption.

In addition to furthering our understanding of the mechanisms underlying statistical associations between genes and AUBs, the results from this study indicate that a relatively high level of accuracy in predicting AUBs can be achieved when combining genetic indicators and intermediate traits, with models explaining 12-36% of the between-person variability in observed AUBs. While genetic indices such as PGSs may eventually be helpful for precision medicine, combining these with information on other risk and protective factors might also be an immediately applicable way to make interventions more effective. More specifically, while the (direct) effects of the PGSs were low, they could be used to identify individuals with higher genetic risk and target cognitive interventions towards the most relevant intermediate mechanisms, such as reshaping drinking motives and/or thoughts about the rewarding effects of alcohol. Interventions targeted towards individuals with a low alcohol sensitivity have already been shown to be useful (Savage et al., 2015), and it is even possible to use self-reports of genetically-influenced traits like alcohol sensitivity when selecting on measured genes is not possible or not desired. Large-scale investigations of specific AUBs are needed to continue refining their genetic etiology and lead to better personalization of such interventions. Furthermore, a systematic application and evaluation is needed to determine whether personalized interventions based on genetic predispositions can improve upon the efficacy of existing programs (Strøm et al., 2014).

While promising, this study’s results should be interpreted in the context of several limitations. The sample was representative of the student population from which it was derived (Dick et al., 2014), but not necessarily generalizable beyond university students or outside of the European subgroup that the sample was restricted to for genetic analyses.

Attrition across waves could reduce this representativeness further, although the use of measures from early waves and aggregate measures across available waves attenuates this concern somewhat. Causal inferences are also not certain; although PGSs based on DNA can reasonably be assumed to come before all other measures, the direction of causation cannot be fully resolved between mediators and outcomes. Most mediators capture either stable traits (personality) or measures prior to/soon after drinking initiation (alcohol sensitivity; alcohol expectancies), but the possibility of reverse causality cannot be ruled out. It has also been demonstrated that the nature of the relationship between drinking motives and AUBs changes during the transition into young adulthood (Savage and Dick, 2023a), complicating the question of whether motives mediate the relationship between genes and AUBs or vice versa. Finally, while we derived PGSs from the largest available GWAS results for all measures, there is a discrepancy in statistical power between the latent factor AUBs and DPW/AUD GWASs, making comparisons for questions about broad versus specific AUB dimensions difficult. In general, PGSs are likely to be highly underpowered in capturing genetic influences relevant to AUBs and their intermediate causes (Lewis and Vassos, 2020).

In conclusion, this study highlights drinking motives, alcohol sensitivity, and, to a lesser extent, personality traits and alcohol expectancies as important mediators of the genetic influences on multiple dimensions of AUBs. Genetic indices of broad constructs such as total drinks per week or AUD diagnoses are relevant to predicting individual-level AUBs, as are the genetic indices of some more specific AUBs involving beverage preference and dynamic consumption patterns. Additional genetic investigations of diverse AUB dimensions are needed to better understand the processes most relevant for heterogeneous groups of individuals. Further, constructs such as drinking motives and alcohol sensitivity are critical mechanisms which require further attention as endophenotypes for AUBs, especially since they are self-report measures that are fairly easy to collect at scale. Combining genetic information with such intermediate processes has the potential to drive forward improved and personalized prevention and treatment applications.

## Data Availability

Data from this study are available to qualified researchers via dbGaP (phs001754.v4.p2) or via to qualified researchers who provide the appropriate signed data use agreement.

## Acknowledgements

This research was funded by a grant to J.E.S. (VI.VENI.201G-064) from The Netherlands Organization for Scientific Research (NWO). Spit for Science has been supported by Virginia Commonwealth University, P20AA017828, R37AA011408, K02AA018755, P50AA022537, and K01AA024152 from the National Institute on Alcohol Abuse and Alcoholism, UL1RR031990 from the National Center for Research Resources and National Institutes of Health Roadmap for Medical Research, as well as support by the Center for the Study of Tobacco Products at VCU. REDCap support provided by CTSA award UM1TR004360 from the National Center for Advancing Translational Sciences. The content is solely the responsibility of the authors and does not necessarily represent the views of the respective funding agencies. The funding agencies had no role in the study design, data analysis, manuscript preparation, or decision to submit for publication. We would like to thank Dr. Danielle Dick for founding and directing the Spit for Science Registry from 2011- 2022, and the Spit for Science participants for making this study a success, as well as the many University faculty, students, and staff who contributed to the design and implementation of the project. Secondary analyses included data from the Million Veteran Program, Office of Research and Development, Veterans Health Administration, and was supported by the Veterans Administration (VA) Million Veteran Program (MVP). The authors thank the staff, researchers, and volunteers, who have contributed to MVP, and especially participants who previously served their country in the military and now generously agreed to enroll in the study. (See https://www.research.va.gov/mvp/ for more details).

## References

Adams, Z. W., Kaiser, A. J., Lynam, D. R., Charnigo, R. J. & Milich, R. 2012. Drinking motives as mediators of the impulsivity-substance use relation: pathways for negative urgency, lack of premeditation, and sensation seeking. Addict Behav, 37, 848–55.

Agrawal, A., Dick, D. M., Bucholz, K. K., Madden, P. A., Cooper, M. L., Sher, K. J. & Heath, A. C. 2008. Drinking expectancies and motives: a genetic study of young adult women. Addiction, 103, 194–204.

Agrawal, A., Freedman, N. D., Cheng, Y. C., Lin, P., Shaffer, J. R., Sun, Q., Taylor, K., Yaspan, B., Cole, J. W., Cornelis, M. C., Desensi, R. S., Fitzpatrick, A., Heiss, G., Kang, J. H., O’connell, J., Bennett, S., Bookman, E., Bucholz, K. K., Caporaso, N., Crout, R., Dick, D. M., Edenberg, H. J., Goate, A., Hesselbrock, V., Kittner, S., Kramer, J., Nurnberger, J. I., Jr., Qi, L., Rice, J. P., Schuckit, M., Van Dam, R. M., Boerwinkle, E., Hu, F., Levy, S., Marazita, M., Mitchell, B. D., Pasquale, L. R. & Bierut, L. J. 2012. Measuring alcohol consumption for genomic meta-analyses of alcohol intake: opportunities and challenges. Am J Clin Nutr, 95, 539–47.

Bohn, M. J., Babor, T. F. & Kranzler, H. R. 1995. The Alcohol Use Disorders Identification Test (AUDIT): validation of a screening instrument for use in medical settings. Journal of Studies on Alcohol, 56, 423–432.

Bresin, K. & Mekawi, Y. 2021. The "Why" of Drinking Matters: A Meta-Analysis of the Association Between Drinking Motives and Drinking Outcomes. Alcohol Clin Exp Res, 45, 38–50.

Britton, A., Ben-Shlomo, Y., Benzeval, M., Kuh, D. & Bell, S. 2015. Life course trajectories of alcohol consumption in the United Kingdom using longitudinal data from nine cohort studies. BMC Med, 13, 47.

Bucholz, K. K., Cadoret, R., Cloninger, C. R., Dinwiddie, S. H., Hesselbrock, V. M., Nurnberger, J. L., Reich, T., Schmidt, I. & Schuckit, M. A. 1994. A new, semi-structured psychiatric interview for use in genetic linkage studies: A report on the reliability of the SSAGA. Journal of Studies on Alcohol, 55, 149–158.

Chang, C. C., Chow, C. C., Tellier, L. C., Vattikuti, S., Purcell, S. M. & Lee, J. J. 2015. Second-generation PLINK: rising to the challenge of larger and richer datasets. Gigascience, 4, 7.

Connor, K. M. & Davidson, J. R. 2003. Development of a new resilience scale: the Connor-Davidson Resilience Scale (CD-RISC). Depress Anxiety, 18, 76–82.

Cooper, M. L. 1994. Motivations for alcohol use among adolescents: Development and validation of a four-factor model. Psychological Assessment, 6, 117–128.

Cronce, J. M. & Larimer, M. E. 2011. Individual-focused approaches to the prevention of college student drinking. Alcohol Res Health, 34, 210–21.

Deak, J. D. & Johnson, E. C. 2021. Genetics of substance use disorders: a review. Psychological Medicine, 51, 2189–2200.

Dick, D. M., Meyers, J. L., Rose, R. J., Kaprio, J. & Kendler, K. S. 2011. Measures of current alcohol consumption and problems: two independent twin studies suggest a complex genetic architecture. Alcoholism: Clinical & Experimental Research, 35, 2152–61.

Dick, D. M., Nasim, A., Edwards, A. C., Salvatore, J. E., Cho, S. B., Adkins, A., Meyers, J., Yan, J., Cooke, M., Clifford, J., Goyal, N., Halberstadt, L., Ailstock, K., Neale, Z., Opalesky, J., Hancock, L., Donovan, K. K., Sun, C., Riley, B. & Kendler, K. S. 2014. Spit for Science: launching a longitudinal study of genetic and environmental influences on substance use and emotional health at a large US university. Front Genet, 5, 47.

Fromme, K., Stroot, E. A. & Kaplan, D. 1993. Comprehensive effects of alcohol: Development and psychometric assessment of a new expectancy questionnaire. Psychological Assessment, 5, 19–26.

Ge, T., Chen, C. Y., Ni, Y., Feng, Y. A. & Smoller, J. W. 2019. Polygenic prediction via Bayesian regression and continuous shrinkage priors. Nat Commun, 10, 1776.

Gunn, R. L., Finn, P. R., Endres, M. J., Gerst, K. R. & Spinola, S. 2013. Dimensions of disinhibited personality and their relation with alcohol use and problems. Addict Behav, 38, 2352–60.

Harris, P. A., Taylor, R., Thielke, R., Payne, J., Gonzalez, N. & Conde, J. G. 2009. Research electronic data capture (REDCap)--a metadata-driven methodology and workflow process for providing translational research informatics support. J Biomed Inform, 42, 377–81.

Hingson, R. W. 2010. Focus on: College drinking and related problems: magnitude and prevention of college drinking and related problems. Alcohol Res Health, 33, 45–54.

Holton, A., Boland, F., Gallagher, P., Fahey, T., Kenny, R. & Cousins, G. 2019. Life Course Transitions and Changes in Alcohol Consumption Among Older Irish Adults: Results From The Irish Longitudinal Study on Ageing (TILDA). J Aging Health, 31, 1568–1588.

John, O. P. & Srivastava, S. 1999. The Big-Five trait taxonomy: History, measurement, and theoretical perspectives. In: Pervin, L. A. & John, O. P. (eds.) Handbook of personality: Theory and research, Vol. 2, (102–138). New York: Guilford Press.

Kendler, K. S., Gardner, C. & Dick, D. M. 2011. Predicting alcohol consumption in adolescence from alcohol-specific and general externalizing genetic risk factors, key environmental exposures and their interaction. Psychol Med, 41, 1507–16.

Kendler, K. S., Ohlsson, H., Edwards, A. C., Sundquist, J. & Sundquist, K. 2021. Mediational Pathways From Genetic Risk to Alcohol Use Disorder in Swedish Men and Women. J Stud Alcohol Drugs, 82, 431–438.

Kuntsche, E., Knibbe, R., Gmel, G. & Engels, R. 2005. Why do young people drink? A review of drinking motives. Clinical Psychology Review, 25, 841–861.

Lai, D., Wetherill, L., Kapoor, M., Johnson, E. C., Schwandt, M., Ramchandani, V. A., Goldman, D., Joslyn, G., Rao, X., Liu, Y., Farris, S., Mayfield, R. D., Dick, D., Hesselbrock, V., Kramer, J., Mccutcheon, V. V., Nurnberger, J., Tischfield, J., Goate, A., Edenberg, H. J., Porjesz, B., Agrawal, A., Foroud, T. & Schuckit, M. 2020. Genome-wide association studies of the self-rating of effects of ethanol (SRE). Addict Biol, 25, e12800.

Leggat, G., Livingston, M., Kuntsche, S. & Callinan, S. 2022. Alcohol consumption trajectories over the Australian life course. Addiction, 117, 1931–1939.

Lewis, C. M. & Vassos, E. 2020. Polygenic risk scores: from research tools to clinical instruments. Genome Med, 12, 44.

Li, J. J., Savage, J. E., Kendler, K. S., Hickman, M., Mahedy, L., Macleod, J., Kaprio, J., Rose, R. J. & Dick, D. M. 2017. Polygenic Risk, Personality Dimensions, and Adolescent Alcohol Use Problems: A Longitudinal Study. J Stud Alcohol Drugs, 78, 442–451.

Litten, R. Z., Ryan, M. L., Falk, D. E., Reilly, M., Fertig, J. B. & Koob, G. F. 2015. Heterogeneity of Alcohol Use Disorder: Understanding Mechanisms to Advance Personalized Treatment. Alcoholism: Clinical and Experimental Research, 39, 579–584.

Littlefield, A. K., Agrawal, A., Ellingson, J. M., Kristjansson, S., Madden, P. A. F., Bucholz, K. K., Slutske, W. S., Heath, A. C. & Sher, K. J. 2011. Does Variance in Drinking Motives Explain the Genetic Overlap Between Personality and Alcohol Use Disorder Symptoms? A Twin Study of Young Women. Alcoholism: Clinical and Experimental Research, 35, 2242–2250.

Lynam, D. R., Smith, G. T., Whiteside, S. P. & Cydera, M. A. 2006. The UPPS-P: Assessing five personality pathways to impulsive behavior. West Lafayette, IN: Purdue University.

Mallard, T. T., Savage, J. E., Johnson, E. C., Huang, Y., Edwards, A. C., Hottenga, J. J., Grotzinger, A. D., Gustavson, D. E., Jennings, M. V., Anokhin, A., Dick, D. M., Edenberg, H. J., Kramer, J. R., Lai, D., Meyers, J. L., Pandey, A. K., Harden, K. P., Nivard, M. G., DE Geus, E. J. C., Boomsma, D. I., Agrawal, A., Davis, L. K., Clarke, T. K., Palmer, A. A. & Sanchez-Roige, S. 2022. Item-Level Genome-Wide Association Study of the Alcohol Use Disorders Identification Test in Three Population-Based Cohorts. Am J Psychiatry, 179, 58–70.

Muthén, L. K. & Muthén, B. O. 2011. Mplus User’s Guide *(Seventh Edition)*, Los Angeles, Ca, Muthén & Muthén.

Peterson, R. E., Edwards, A. C., Bacanu, S. A., Dick, D. M., Kendler, K. S. & Webb, B. T. 2017. The utility of empirically assigning ancestry groups in cross-population genetic studies of addiction. Am J Addict, 26, 494–501.

Poore, H. E., Hatoum, A., Mallard, T. T., Sanchez-Roige, S., Waldman, I. D., Palmer, A. A., Harden, K. P., Barr, P. B. & Dick, D. M. 2023. A multivariate approach to understanding the genetic overlap between externalizing phenotypes and substance use disorders. Addiction Biology, 28, e13319.

Prescott, C. A., Cross, R. J., Kuhn, J. W., Horn, J. L. & Kendler, K. S. 2004. Is risk for alcoholism mediated by individual differences in drinking motivations? Alcohol Clin Exp Res, 28, 29–39.

R Core Team 2017. R: A language and environment for statistical computing. Vienna, Austria: R Foundation by Statistical Computing.

Rehm, J., Rehn, N., Room, R., Monteiro, M., Gmel, G., Jernigan, D. & Frick, U. 2003. The global distribution of average volume of alcohol consumption and patterns of drinking. Eur Addict Res, 9, 147–56.

Salvatore, J. E., Aliev, F., Edwards, A. C., Evans, D. M., Macleod, J., Hickman, M., Lewis, G., Kendler, K. S., Loukola, A., Korhonen, T., Latvala, A., Rose, R. J., Kaprio, J. & Dick, D. M. 2014. Polygenic Scores Predict Alcohol Problems in an Independent Sample and Show Moderation by the Environment. Genes, 5, 330–46.

Salvatore, J. E., Thomas, N. S., Cho, S. B., Adkins, A., Kendler, K. S. & Dick, D. M. 2016. The role of romantic relationship status in pathways of risk for emerging adult alcohol use. Psychol Addict Behav, 30, 335–44.

Sanchez-Roige, S., Palmer, A. A., Fontanillas, P., Elson, S. L., Adams, M. J., Howard, D. M., Edenberg, H. J., Davies, G., Crist, R. C., Deary, I. J., Mcintosh, A. M. & Clarke, T.-K. 2019. Genome-Wide Association Study Meta-Analysis of the Alcohol Use Disorders Identification Test (AUDIT) in Two Population-Based Cohorts. American Journal of Psychiatry, 176, 107–118.

Saunders, G. R. B., Wang, X., Chen, F., Jang, S. K., Liu, M., Wang, C., Gao, S., Jiang, Y., Khunsriraksakul, C., Otto, J. M., Addison, C., Akiyama, M., Albert, C. M., Aliev, F., Alonso, A., Arnett, D. K., Ashley-Koch, A. E., Ashrani, A. A., Barnes, K. C., Barr, R. G., Bartz, T. M., Becker, D. M., Bielak, L. F., Benjamin, E. J., Bis, J. C., Bjornsdottir, G., Blangero, J., Bleecker, E. R., Boardman, J. D., Boerwinkle, E., Boomsma, D. I., Boorgula, M. P., Bowden, D. W., Brody, J. A., Cade, B. E., Chasman, D. I., Chavan, S., Chen, Y. I., Chen, Z., Cheng, I., Cho, M. H., Choquet, H., Cole, J. W., Cornelis, M. C., Cucca, F., Curran, J. E., De Andrade, M., Dick, D. M., Docherty, A. R., Duggirala, R., Eaton, C. B., Ehringer, M. A., Esko, T., Faul, J. D., Fernandes Silva, L., Fiorillo, E., Fornage, M., Freedman, B. I., Gabrielsen, M. E., Garrett, M. E., Gharib, S. A., Gieger, C., Gillespie, N., Glahn, D. C., Gordon, S. D., Gu, C. C., Gu, D., Gudbjartsson, D. F., Guo, X., Haessler, J., Hall, M. E., Haller, T., Harris, K. M., He, J., Herd, P., Hewitt, J. K., Hickie, I., Hidalgo, B., Hokanson, J. E., Hopfer, C., Hottenga, J., Hou, L., Huang, H., Hung, Y. J., Hunter, D. J., Hveem, K., Hwang, S. J., Hwu, C. M., Iacono, W., Irvin, M. R., Jee, Y. H., Johnson, E. O., Joo, Y. Y., Jorgenson, E., Justice, A. E., Kamatani, Y., Kaplan, R. C., Kaprio, J., Kardia, S. L. R., Keller, M. C., et al. 2022. Genetic diversity fuels gene discovery for tobacco and alcohol use. Nature, 612, 720–724.

Savage, J. E., Barr, P. B., Phung, T., Lee, Y. H., Yingzhe, Z., Investigators, C., Ge, T., Smoller, J. W., Davis, L. K., Meyers, J., Porjesz, B., Posthuma, D., Mallard, T. T. & Sanchez-Roige, S. 2023. Genetic Heterogeneity Across Dimensions of Alcohol Use Behaviors. medRxiv, 2023.12.26.23300537.

Savage, J. E. & Dick, D. M. 2023a. Drinking Motives, Alcohol Misuse, and Internalizing and Externalizing Psychopathology across College: A Cross-Lagged Panel Study. Subst Use Misuse, 58, 1377–1387.

Savage, J. E. & Dick, D. M. 2023b. Internalizing and externalizing subtypes of alcohol misuse and their relation to drinking motives. Addict Behav, 136, 107461.

Savage, J. E., Neale, Z., Cho, S. B., Hancock, L., Kalmijn, J. A., Smith, T. L., Schuckit, M. A., Donovan, K. K. & Dick, D. M. 2015. Level of response to alcohol as a factor for targeted prevention in college students. Alcohol Clin Exp Res, 39, 2215–23.

Savage, J. E., Peterson, R. E., Aliev, F. & Dick, D. M. 2022. Genetic and environmental etiology of drinking motives in college students. Alcohol Clin Exp Res, 46, 1783–1796.

Schuckit, M. A. 2009. An overview of genetic influences in alcoholism. Journal of substance abuse treatment, 36, 5.

Schuckit, M. A., Smith, T. L. & Tipp, J. E. 1997. The Self-Rating of the Effects of alcohol (SRE) form as a retrospective measure of the risk for alcoholism. Addiction, 92, 979–88.

Sieri, S., Agudo, A., Kesse, E., Klipstein-Grobusch, K., San-José, B., Welch, A. A., Krogh, V., Luben, R., Allen, N., Overvad, K., Tjønneland, A., Clavel-Chapelon, F., Thiébaut, A., Miller, A. B., Boeing, H., Kolyva, M., Saieva, C., Celentano, E., Ocké, M. C., Peeters, P. H., Brustad, M., Kumle, M., Dorronsoro, M., Fernandez Feito, A., Mattisson, I., Weinehall, L., Riboli, E. & Slimani, N. 2002. Patterns of alcohol consumption in 10 European countries participating in the European Prospective Investigation into Cancer and Nutrition (EPIC) project. Public Health Nutr, 5, 1287–96.

Simpura, J. & Karlsson, T. 2001. Trends in drinking patterns among adult population in 15 European countries, 1950 to 2000: a review. Nordic Studies on Alcohol and Drugs, 18, 31-53.

Sliedrecht, W., De Waart, R., Witkiewitz, K. & Roozen, H. G. 2019. Alcohol use disorder relapse factors: A systematic review. Psychiatry Res, 278, 97–115.

Stautz, K. & Cooper, A. 2013. Impulsivity-related personality traits and adolescent alcohol use: a meta-analytic review. Clin Psychol Rev, 33, 574–92.

Strøm, H. K., Adolfsen, F., Fossum, S., Kaiser, S. & Martinussen, M. 2014. Effectiveness of school-based preventive interventions on adolescent alcohol use: a meta-analysis of randomized controlled trials. Subst Abuse Treat Prev Policy, 9, 48.

Verhulst, B., Neale, M. C. & Kendler, K. S. 2015. The heritability of alcohol use disorders: a meta-analysis of twin and adoption studies. Psychol Med, 45, 1061–72.

Webb, B. T., Edwards, A. C., Wolen, A. R., Salvatore, J. E., Aliev, F., Riley, B. P., Sun, C., Williamson, V. S., Kitchens, J. N., Pedersen, K., Adkins, A., Cooke, M. E., Savage, J. E., Neale, Z., Cho, S. B., Dick, D. M. & Kendler, K. S. 2017. Molecular Genetic Influences on Normative and Problematic Alcohol Use in a Population-Based Sample of College Students. Frontiers in Genetics, 8.

Whelan, R., Watts, R., Orr, C. A., Althoff, R. R., Artiges, E., Banaschewski, T., Barker, G. J., Bokde, A. L., Buchel, C., Carvalho, F. M., Conrod, P. J., Flor, H., Fauth-Buhler, M., Frouin, V., Gallinat, J., Gan, G., Gowland, P., Heinz, A., Ittermann, B., Lawrence, C., Mann, K., Martinot, J. L., Nees, F., Ortiz, N., Paillere-Martinot, M. L., Paus, T., Pausova, Z., Rietschel, M., Robbins, T. W., Smolka, M. N., Strohle, A., Schumann, G. & Garavan, H. 2014. Neuropsychosocial profiles of current and future adolescent alcohol misusers. Nature, 512, 185–9.

Wong, C. C. Y. & Schumann, G. 2008. Genetics of addictions: strategies for addressing heterogeneity and polygenicity of substance use disorders. Philosophical Transactions of the Royal Society B: Biological Sciences, 363, 3213–3222.

Zhou, H., Kember, R. L., Deak, J. D., Xu, H., Toikumo, S., Yuan, K., Lind, P. A., Farajzadeh, L., Wang, L., Hatoum, A. S., Johnson, J., Lee, H., Mallard, T. T., Xu, J., Johnston, K. J. A., Johnson, E. C., Nielsen, T. T., Galimberti, M., Dao, C., Levey, D. F., Overstreet, C., Byrne, E. M., Gillespie, N. A., Gordon, S., Hickie, I. B., Whitfield, J. B., Xu, K., Zhao, H., Huckins, L. M., Davis, L. K., Sanchez-Roige, S., Madden, P. A. F., Heath, A. C., Medland, S. E., Martin, N. G., Ge, T., Smoller, J. W., Hougaard, D. M., Børglum, A. D., Demontis, D., Krystal, J. H., Gaziano, J. M., Edenberg, H. J., Agrawal, A., Justice, A. C., Stein, M. B., Kranzler, H. R. & Gelernter, J. 2023. Multi-ancestry study of the genetics of problematic alcohol use in over 1 million individuals. Nat Med, 29, 3184–3192.

Zhou, H., Sealock, J. M., Sanchez-Roige, S., Clarke, T.-K., Levey, D. F., Cheng, Z., Li, B., Polimanti, R., Kember, R. L., Smith, R. V., Thygesen, J. H., Morgan, M. Y., Atkinson, S. R., Thursz, M. R., Nyegaard, M., Mattheisen, M., Børglum, A. D., Johnson, E. C., Justice, A. C., Palmer, A. A., Mcquillin, A., Davis, L. K., Edenberg, H. J., Agrawal, A., Kranzler, H. R. & Gelernter, J. 2020. Genome-wide meta-analysis of problematic alcohol use in 435,563 individuals yields insights into biology and relationships with other traits. Nature neuroscience, 23, 809–818.

